# Development and validation of a novel mobility test for IRDs, from reality to virtual reality

**DOI:** 10.1101/2023.02.01.23285189

**Authors:** Colas Authié, Mylène Poujade, Alireza Talebi, Alexis Defer, Ariel Zenouda, Cécilia Coen, Saddek Mohand-Said, Philippe Chaumet-Riffaud, Isabelle Audo, José-Alain Sahel

**Affiliations:** Streetlab, Paris, 75012 France; Sorbonne Universités, INSERM U968, CNRS UMR7210, Institut de la Vision, Paris, France; Hôpital National de la Vision des Quinze-Vingts, DHU Sight Restore, Centre de Référence Maladies Rares REFERET, INSERM-DHOS CIC 1423, Paris, France; Department of Ophthalmology, School of Medicine, University of Pittsburgh, Pittsburgh, Pennsylvania, United States

**Keywords:** Locomotion, mobility, virtual reality, rod-cone dystrophy, retinitis pigmentosa, locomotion, dim light, performance-based outcome

## Abstract

**Purpose:** To validate a novel mobility test (MOST, MObility Standardized Test) and performance outcomes in real (RL) and virtual (VR) environments to be used for interventional clinical studies in order to characterize vision impairment in rod-cone dystrophies, also known as retinitis pigmentosa (RP).

**Design:** Prospective, interventional, non-invasive, longitudinal study (test-retest).

**Participants:** 89 participants in three experimental phases: 15 non visually impaired (controls) in Phase 1 (average age, 27.4 years; 66% women), 14 participants with RP in Phase 2 (average age, 45.2 years, 36% women), and 60 participants (30 RP; average age, 47.4; 44.6% women; and 30 controls, average age, 47.6 years; 45.4% women) in Phase 3.

**Methods:** We designed a mobility test (MOST) to be used in both VR and RL and ran three experimental studies to (1) validate the difficulty of the mobility courses, (2) determine the optimal number of light levels and training trials, and (3) validate the reproducibility (test-retest), reliability (VR/RL), sensitivity, and construct, and content validity of the test. A comprehensive ophthalmologic examination was performed in all subjects.

**Main outcomes measures:** The primary outcome is the performance score in the mobility test. The secondary outcomes include visual acuity, contrast sensitivity, dark adaptation thresholds, static and kinetic visual field parameters, and ellipsoid zone from optical coherence tomography. Correlation between the performance score in the mobility tests and visual function were assessed.

**Results:** Results revealed that the mobility courses developed exhibited statistically similar difficulty, and that five trials are sufficient to control for the learning effect in a session. MOST is highly reproducible (test-retest intra-class correlations > .98) and reliable (correlation VR/RL = .98). MOST achieved a discrimination between RP participants and controls (accuracy larger than 95% in all conditions) and between early and late stages of the disease (mean accuracy of 82.3%). The performance score is correlated with visual function parameter (.57 to .94).

**Conclusion:** MOST is a tool offering validated mobility test, a controlled learning effect, which demonstrates excellent reproducibility and high agreement between real and virtual conditions, as well as sensitivity and specificity to measure disease progression and therapeutic benefit in IRD.

## Introduction

Inherited retinal diseases (IRD) characterized by photoreceptor loss are a major cause of untreatable blindness(1). The impact of vision loss on quality of life require the development of effective technologies for restoring or protecting vision(2). The approval of Voretigene Neparvovec (VN) for the treatment of IRDs caused by mutations in RPE65(3) gene, marked an important milestone, leading to numerous trials in gene and cell therapy, while prosthetic vision technologies were in development(4). A crucial factor in determining the efficacy of the therapy is the selection of appropriate outcome measures(5–7). In addition to evaluating retinal structure or visual function outcomes, such as visual acuity, contrast sensitivity and visual field, it is key to quantify functional vision, as defined by the patient’s ability to perform vision-dependent tasks that are essential to maintain autonomy(8, 9). This is of particular importance because conventional clinical visual function tests (e.g., visual acuity) do not accurately reflect the visual deficits that patients experience in daily life(10, 11).

Translational researchers have worked to build performance-based outcomes for a variety of activities of daily living, such as orientation and mobility(6, 12–14), corresponding to a major difficulty for IRD patients, especially under low light conditions(15). The approval of VN therapy was based on such metrics, and a multi-luminance mobility test (MLMT) performance was even used as the primary outcome of the phase III VN trial(3). However, the existing tools have several drawbacks that need to be improved: 1) they show poor sensitivity to discriminate between early and advanced forms of the disease; 2) they are not always ecological (e.g., with MLMT, the participants’ natural footsteps are modified small size of the mobility space, which may require a learning phase that is difficult for the experimenter to control and measure); 3) they do not systematically control for learning effects within and between sessions, to verify that the performance improvement is related to the restoration of the functional vision; 4) they do not include a continuous performance score considering both patient’s accuracy and speed during the task; 5) they are very difficult and expensive to deploy, replicate, and standardize for multi-center trials and post-approval validation studies; 6) they require a considerable amount of material, setup, and time for the experimenter and patients. To improve the current tools with an exportable test, the validation of the test in virtual reality (VR) has become essential.

VR is a widely used tool in neuroscience research(16), but also for performance assessment, including attempts in ophthalmology(13, 17–19). VR benefits comprise a total control of experimental parameters (including light level), fast and objective behavioral measurement, reproducibility between multiple assessment centers, and participant safety. Significant work still needs to be done to demonstrate the reproducibility of an outcome from the real world to VR, particularly in a low-vision population. Translational research may help to better understand the ecological validity of VR, either in terms of control of physical parameters (e.g., luminosity(20)) or regarding the sensorimotor behavior(21). Significant work needs to be done to demonstrate outcome reproducibility across real world and VR, particularly in a low vision population. These issues are relevant beyond the field of clinical trials in retinal degenerations, as a behavioral neuroscience question with applications in various fields (e.g., psychiatric disorders(22), post-stroke rehabilitation(23)).

Developing new functional vision outcomes for interventional clinical trials targeting IRD is fundamental for the improvement of therapies and patient monitoring. The objective of this study is to present the different phases of the development of a new mobility outcome (MOST, MObility Standardized Test), and to validate MOST. MOST was developed and studied in real-life environment (RL) and virtual reality environment (VR). This study was elaborated in three phases. Phase 1 was dedicated to measure the performance in MOST of control participants in VR in order to homogenize the difficulty of the mobility courses. In Phase 2 the optimal number of luminance levels and training trials with RP participants (VR and RL) was determined. Finally, in Phase 3, we assessed MOST by measuring the construct and content validity, the reproducibility, and the sensitivity (VR and RL) with RP and control participants.

## Methods

Participants were included in a prospective, interventional, non-invasive, longitudinal study designed to compare the performance of RP patients and control participants in behavioral tasks. Inclusion and screening were conducted at the XV-XX National Ophthalmology Hospital in Paris, France, whereas all behavioral assessments were conducted at Streetlab(6), Paris, France. The study was approved by the Ouest V Ethics Committee (CPP 19.01446.190402-MS03; IDRCB: 2019-A00483-54; ClinicalTrials.gov ID: NCT04448860) in accordance with the Declaration of Helsinki. Written informed consent was obtained from all participants.

The aim of the present study was to validate the use of a mobility performance test (MOST, MObility Standardized Test) conducted out in real life (RL) and in virtual reality (VR) to be used in interventional clinical studies with inherited retinal disease conditions. For this purpose, we first compared the difficulty level of the mobility courses with control participants in VR (Phase 1). Then, we determined the optimal number of light levels and training trials (Phase 2) with RP patients in both VR and RL conditions. Finally, in the validation phase (Phase 3), we evaluated the construct and content validity, the reproducibility and the sensitivity of MOST (in both VR and RL) with RP patients and control participants. Participants involved in the three phases of the study were different, for independent validation purposes.

All participants, except controls in Phase 1, underwent a comprehensive ophthalmologic examination, including the review of medical history, binocular and monocular best-corrected visual acuity and contrast sensitivity, slit lamp biomicroscopy and fundus examination, intraocular pressure measurement, retinophotography, static and kinetic visual fields, binocular dark adaptation, microperimetry (MAIA, Centervue, Padova, Italy), and spectral domain optical coherence tomography (SD-OCT Spectralis, Heidelberg, Germany). Visual acuity was measured using the Early Treatment Diabetic Retinopathy Study (ETDRS) chart with optimal optical correction, and it was expressed as the logarithm of the minimum angle of resolution (logMAR). Contrast sensitivity was measured with the Pelli-Robson chart and expressed in logCS (Haag-Streit, Mason, OH, USA). Static visual field was assessed monocularly with a 24-2 strategy using Octopus® 900 (Haag-Streit, Inc., Köniz, Switzerland) to measure the mean sensitivity and deficit. Goldmann kinetic perimetry assessment was performed in monocular and binocular conditions (III4e, V4e, I4e), and the central island area, total visual field area, horizontal and vertical diameters were collected. In addition, two SD-OCT graders independently delimited the boundaries of the preserved ellipsoid zone (EZ). In order to grade disease severity, we used a classification(24) combining visual acuity, Goldmann visual field diameter, and EZ size. Dark adaptation thresholds were measured binocularly with Metrovision MonPackOne (MetroVision, Perenchies, France) after 5 and 20 minutes of dark adaptation. After completion of the mobility test, participants answered an ad hoc questionnaire to evaluate comfort, usefulness, usability, and perception of danger (Supplementary Table 1, only in Phase 3).

The inclusion criteria common to all participants required being 18 to 75 years old, with no participation in any other clinical trial that may interfere with this study, an independent walking ability, a Mini-Mental State Examination(25) score without visual items ≥ 20*/*25, and a proficient knowledge of the French language to understand the tasks and instructions. RP patients had to have a confirmed diagnosis of retinitis pigmentosa by an ophthalmologist. We included RP patients with varying degrees of visual field, acuity, contrast sensitivity, and electroretinogram anomalies. Control participants had to have a best corrected visual acuity greater than or equal to 20/25, a normal semi-automatic kinetic visual field (except for Phase 1), a normal walking ability, and being aged matched to RP participants (Phase 3). Participants from all groups were excluded if they presented any other ocular or systemic disease that could affect either the optic nerve or the visual field.

## Protocol

For the three phases of the study, participants performed a mobility test in either real life (RL) or virtual reality (VR) conditions, using multiple mobility courses. VR experiments were conducted by using a HTC Vive Pro Eye headset. In Phase 1, controls performed the MOST test in a single VR session, at maximum light intensity, and in binocular condition. After 10 training trials, the participants performed 28 test trials, each one on a different and randomized mobility course (in both training and test phases). We analyzed performance to determine if difficulty between mobility courses were comparable, and usable in later phases. In Phase 2, RP patients performed four test sessions, in VR and RL, the first day of the study (D1) and one month later (M1). During each session, after an explanation of the instructions and a demonstration trial, patients performed 10 training trials followed by a 20-minute dark adaptation phase (at 1 lux for RL, and at the lowest light level of the VR headset for VR, see Appendix 1). Afterward, they performed 14 test trials, each one on a different and randomized mobility course, and under a different luminance level (from 1 to 400 lux). The performance of the patients was then analyzed to determine the number of training trials and illumination levels required for the next phase. In Phase 3, age-matched RP participants and controls performed four test sessions (D1/M1, VR/RL). Each session included 5 training trials, followed by 20 min of dark adaptation and 18 test trials, with 6 light levels from dim to bright, each one performed in monocular (left and right) and binocular conditions. The mobility course configuration was randomized.

## MOST mobility courses

A total of 38 unique mobility course configurations were used for both RL and VR versions of the test (Supplementary Figure 1). The courses were presented in a rectangular area of 5.2 meters by 3.6 meters, delimited by strips on the ground (Figure 1.A). Each course had the same length (22 m), number of turns (9), and number and type of obstacles (Supplementary Figure 2). Participants were instructed to follow a unique path (60 cm wide) through a maze on their own, to a goal displayed on the ground (gray square of fabric). This path was formed by foldable doors supported by low columns (l20*L20*H74 cm) that meshed the space, and by obstacles blocking the path. On their way, participants were instructed to step over two steps (10 cm high, 50 cm wide) and to duck under two flags (the lower part being at eye level, Supplementary Figure 2). The course also included a dead end (80 cm long), a cone and two high columns (l20*L20*H200 cm) closing the path. Mobility courses were randomly assigned to each trial to avoid any learning effect. In order to measure their maximum performance, participants were instructed to walk as fast as possible, while making as few errors as possible in terms of the course requirements (stepping over the steps and ducking under the flags). The mobility course was strictly identical in VR and RL conditions (Figure 1.B-D). In the RL condition, participants were guided to the starting location with their eyes closed, and they were instructed to open their eyes and start the trial at the sound of an auditory signal. In the VR condition, they reached the starting point – the only visible element in the scene at that point – and they started the test as soon as the maze appeared (Supplementary Video 1). For each training or test trial, we measured the duration of the trial, the number of collisions with the objects constituting the path (doors, low and high columns, cone), the number of steps and flags touched, entries in the dead end and interventions. An intervention was triggered whenever a participant went out of the mobility course, or took the path in the wrong direction (turn-around). All these variables were determined automatically in VR and manually in RL (except for the trial duration). In VR, to compensate for the lack of haptic feedback, each error triggered a specific sound. As VR environments have the potential to induce motion sickness when visual movement does not match physical movement(17), participants had to physically move to navigate in the virtual environment (Figure 1.B). This provides an increased ecological validity, as actual motion is essential when studying mobility and/or wayfinding(26) - which is often overlooked in other VR paradigms.

**Fig. 1.**
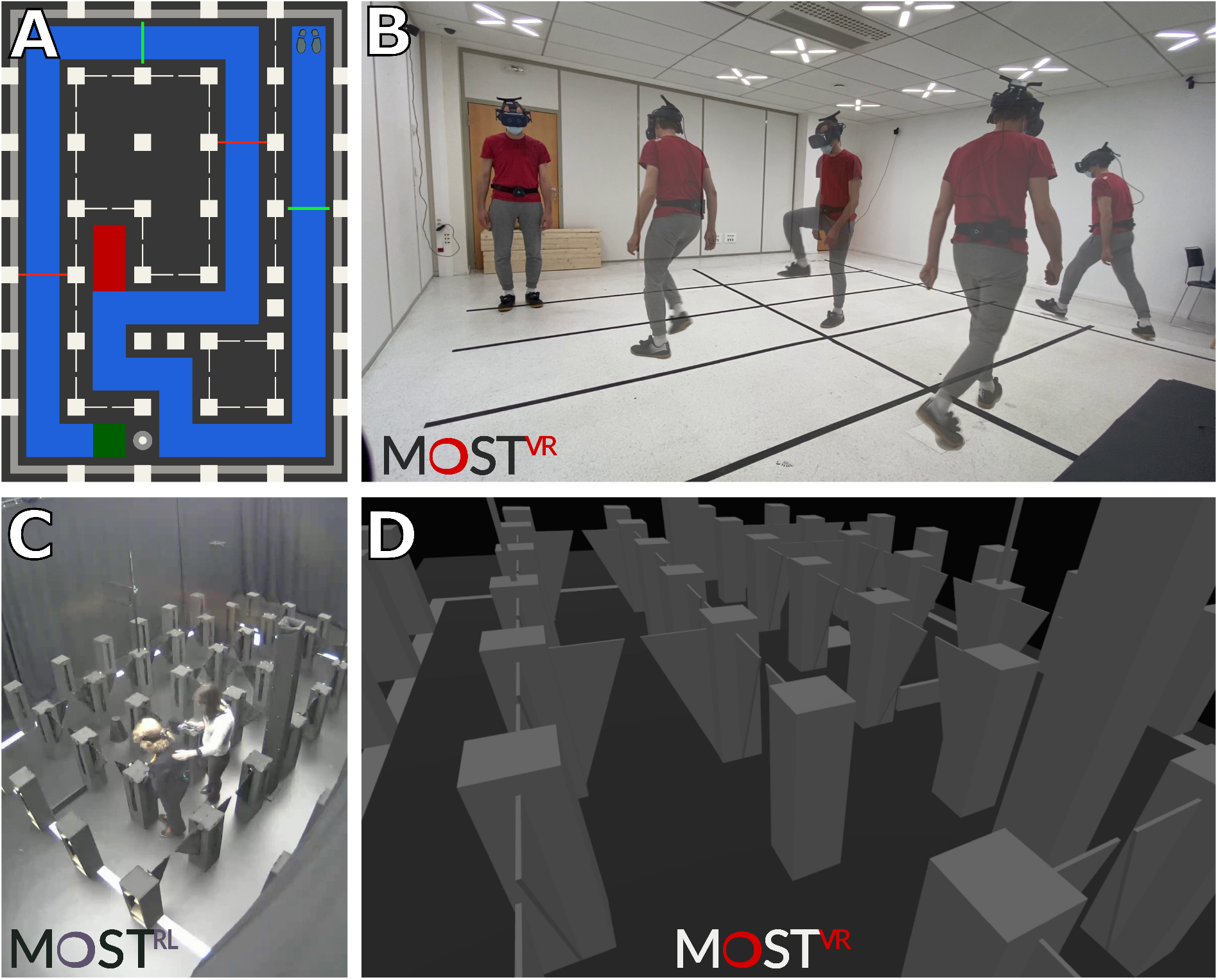
Description of the mobility test (MOST - MObility Standardized Test). MOST was designed to measure difficulties in the daily life of patients with visual impairment. Participants performed the test both in real conditions (physical maze in the artificial street - MOST-RL [**C**]) and in a VR HMD (head mounted display; physical movement in virtual mazes, MOST-VR [B/D]). **A**. Top view. Participants walk through a maze delimited by strips on the ground (gray), forming a rectangle of 5.2 meters by 3.6 meters. Participants must autonomously join the arrival on the ground by following a unique path (highlighted in blue here) leading to a goal (green). This path is formed by folding doors supported by low columns that mesh the space, and by obstacles closing the path. On their way, participants will have to step over two steps (green lines) and to bend down under two flags (red lines). **B**. In MOST-VR, participants move both in the physical space of the laboratory and in a virtual environment. **C**. External view of a participant in MOST-RL (physical locomotion course). **D**. First-person-view of the maze in VR-HMD.

### Luminance levels

RL tests were conducted in a room equipped with a lighting system capable of providing multiple luminance levels ranging from 1 to 400 lux (constant color temperature of 4,000K), to approximate common, real-world lighting levels(12). Luminance was chosen to be evenly distributed in log units(27), across a 14-lux scale in Phase 2 (i.e., 1, 1.6, 2.5, 4, 6.3, 10, 16, 25, 40, 63, 100, 159, 252 and 400 lux), to determine the number of illumination levels required for the test, and a 6-lux scale in Phase 3 (i.e., 1, 3.3, 11, 36, 121 and 400 lux, see Appendix 1). Illuminance was controlled in intensity and color temperature by nine LED panels on the ceiling, and it was measured to be stable over nine measurement locations in the walking area (average variation of 3%, lux meter Chroma Meter CL-200A, Konica Minolta, Tokyo, Japan), and significantly different between all luminance levels. Luminance levels in VR were chosen empirically, but followed the same logic (see Appendix 1). The minimal luminance level was defined as the lowest light condition for which a normally-sighted participant was able to perform the mobility test, and the maximum level corresponded to a light level that visually matched the 400 lux condition in the real environment (i.e., the maximum light level in RL), without glare. As in the RL condition, illumination was measured in VR and luminance levels were evenly distributed in log lux units. The lowest light level in VR was increased between Phase 2 and Phase 3 to better match the performance of patients in VR and RL conditions (Appendix 1).

### Recording apparatus

In the RL condition, two HTC Vive trackers (HTC Corp., New Taipei, Taiwan) were used to record the position of the pelvis and the head of participants. The pelvis position was used to automatically measure trial duration. Mobility errors were coded by an experimenter on-the-fly by using a remote controller. Videos were recorded by video-surveillance cameras to double-check each recorded error offline. In the VR conditions, four HTC Vive trackers were used to track the position of the pelvis and the feet, as the wireless version of the HTC Vive head mounted display (HMD) was used to track the head. The HMD was composed of two AMOLED screens covering a diagonal of 110 degrees of field of view (resolution of 2,880 × 1,600 pixels at 90 Hz). Custom software developed in Unity game engine (2018.2.17f1 version in Phase 2, 2019.3.15f1 in Phase 3, Unity Technologies, San Francisco, CA, USA) recorded the kinematic data from Vive Tackers (RL & VR), triggered the sound system (RL & VR), controlled the lighting system and the video-surveillance cameras (RL), and displayed the virtual environment in the HMD (VR).

### Scoring system

By a custom software developed in Python 3.9.6 (http://www.python.org), we designed a quantitative performance score that combined trial duration and mobility errors. This score ranged from 0 (inability to achieve a trial within a time limit of 160 seconds) to 100 (no errors and duration < 22 seconds). The score of each trial was calculated as a linear combination of a series of sub-scores (Eq. 1):

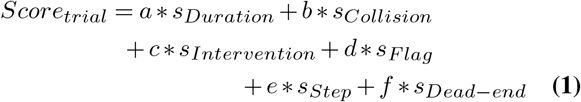

Each sub-score ranged from 0 (minimum performance) to 1 (maximum performance). For example, the sDead-end subscore was equal to 1 if a participant avoided the dead-end, and 0 otherwise; the subscore sStep was equal to 1, 0.5, and 0 for 0-step error, 1-step error, and 2-step errors, respectively. The minimum sub-score sCollision and sIntervention was set to 0 for 10 collisions/interventions, and the minimal and maximal sub-score sDuration was achieved for trial duration of 22 and 160 seconds, respectively. These cutoffs were derived from the distributions of the phase 2 and 3 data in our study. For example, 50% of the control participants’ trials lasted less than 22 seconds, whereas 90% of the patients’ trials lasted less than 122 seconds. Coefficients a to f were empirically determined from experimental constraints and instruction: a = 50, b = 20, c = 20, d = 4, e = 4, f = 2. Because we asked participants to walk as fast as possible making as few errors as possible, we assumed that the duration sub-score (sDuration) should have the same weight (a) as all other sub-scores’ weights (b to f) combined. The other coefficients were determined according to the relative frequency of occurrence of each event (a single dead end, two flags and steps, 9 turns that could lead to collisions and interventions). The score associated to a session was simply taken as the average of the scores of all trials in the session.

### Statistical analysis

Statistical analysis was carried out in R 4.2.2 (http://www.R-project.org). The statistical significance level was set to 0.05. Repeated measures analysis of variance (ANOVA, type II error) or Welch’s t-test were performed in order to assess the effect of the group (RP, control), session (D1, M1), condition (RL, VR), and luminance levels. Tukey’s HSD tests were used for post-hoc analysis whenever necessary. For non-normally distributed variables, Wilcoxon and Mann–Whitney tests with false-discovery rate (FDR) corrections for multiple comparisons were applied. Fisher’s exact test was used for group comparison for categorical variables. Agreement between sessions (i.e., repeatability) and conditions (i.e., reliability) were assessed with intra-class correlation(28) (ICC), mean difference, and 95% limits of agreement from Bland-Altman(29) plots. Relations between the performance score and the visual variables were assessed with Pearson Product-Moment correlation, using FDR correction. Partial eta squared (ηp2) was used to indicate effect size.

### Results

A population of 89 participants were recruited in the three independent experimental phases of the study: 15 healthy volunteers (controls) to validate the difficulty of the mobility courses (Phase 1), 14 RP patients to determine the optimal number of luminance levels and training trials (Phase 2), and 60 participants (30 RP patients and 30 controls) validate the finding of Phase 2 (Phase 3). The demographic and clinical characteristics of the study participants are summarized in Table1.

**Table 1.** Demographic and clinical characteristics of participants included in the three phases of the study. Participants involved in the three phases of the study were all different. Values are presented as mean ± standard deviation (minimum to maximum). *Student t-test. †Fisher exact test. ‡Wilcoxon rank-sum test. BE: Best Eye. Bino.: Binocular.

| Characteristics | Phase 1 | Phase 2 | Phase 3 |  | P Value |
| --- | --- | --- | --- | --- | --- |
|  | Controls<br>(n = 15) | Retinitis pigmentosa<br>(n = 14) | Controls<br>(n = 30) | Retinitis pigmentosa<br>(n = 30) |  |
| Age (yrs) | 27.40 ± 5.18<br>(22 to 40) | 45.28 ± 12.42<br>(29 to 68) | 45.37 ± 14.69<br>(20 to 67) | 44.57 ± 13.31<br>(19 to 65) | 0.82* |
| Gender (% female) | 66.67 | 35.71 | 46.67 | 43.33 | 1.00† |
| Visual Acuity<br>(binocular, logMAR) | -0.15 ± 0.11<br>(-0.30 to 0.00) | 0.18 ± 0.24<br>(-0.08 to 0.84) | -0.14 ± 0.08<br>(-0.30 to -0.04) | 0.24 ± 0.27<br>(-0.22 to 0.84) | <0.001‡ |
| Contrast Sensitivity<br>(binocular, logCS) | - | 1.68 ± 0.18<br>(1.35 to 1.95) | 1.95 ± 0.05<br>(1.80 to 2.10) | 1.39 ± 0.49<br>(0.15 to 2.25) | <0.001‡ |
| Mean Sensitivity<br>(Octopus, BE, dB) | - | 4.32 ± 4.57<br>(0.40 to 18.70) | 25.31 ± 2.31<br>(17.80 to 29.50) | 4.42 ± 4.12<br>(0.41 to 18.14) | <0.001‡ |
| Mean Defect<br>(Octopus, BE, dB) | - | 23.34 ± 3.75<br>(11.90 to 27.10) | 1.94 ± 2.02<br>(-1.30 to 8.20) | 23.03 ± 3.84<br>(10.50 to 27.40) | <0.001‡ |
| Horizontal diameter III4<br>(Goldmann, Bino., °) | - | 50.42 ± 58.04<br>(8.33 to 172.22) | 168.70 ± 8.33<br>(145.55 to 178.89) | 47.45 ± 49.89<br>(4.44 to 170.59) | <0.001‡ |
| Dark Adaptometry Threshold<br>(Metrovision, 20 min., dB) | - | 37.25 ± 10.51<br>(21.00 to 52.00) | 58.84 ± 4.77<br>(51.00 to 67.00) | 32.37 ± 13.81<br>(12.00 to 58.00) | <0.001* |

### Phase 1 - Validating the difficulty levels across MOST mobility courses

First, we compared the difficulty of 28 different mobility courses with control participants (see Supplementary Figure 1 for some examples) in the VR condition only. All performance variables showed no effect of the type of mobility course (all p>.2, see Supplementary Table 2): trial duration, number of collisions, number of errors for dead-end, steps, and errors and flags, and number of interventions by the experimenter to redirect the participant. The mobility courses were therefore of comparable difficulty, and they were used in the following study phases.

### Phase 2 - MOST number of luminance levels and control of learning effects

Second, we tested 14 RP patients in 4 MOST sessions: under RL and VR conditions, the first day (D1), and 1 month after (M1). Patients underwent 10 training trials with the highest luminance level. Then, after a 20-min dark adaptation period, they performed 14 test trials, from the lowest to the highest luminance, while viewing binocularly. All patients were able to perform the test in both RL and VR conditions, but with variable levels of performance. Their performance score improved slightly across training trials: a Wilcoxon signed-rank test revealed that performance was significantly lower in the 1st (Mdn = 88.2%, z = -3.67, p = .002) and 2nd trial (Mdn = 91.1%, z = -2.71, p = .002) as compared to the 10th trial (Mdn = 92.4%). No significant differences were observed between other training trials. These results were determined by large inter-individual variations in terms of behavioral performance. We therefore identified two groups: the slow learners (n = 5) and the fast learners (n = 9, see Figure 2.A). In D1, fast learners need only 1 trial to reach peak performance, in both RL and VR conditions. By contrast, slow learners needed until the 5th trial to learn the task. One month later (M1), the performance of both groups was similar, in both RL and VR, and no more learning was observed. These findings indicated that 5 trials were sufficient to control for a learning effect in this test.

**Fig. 2.**
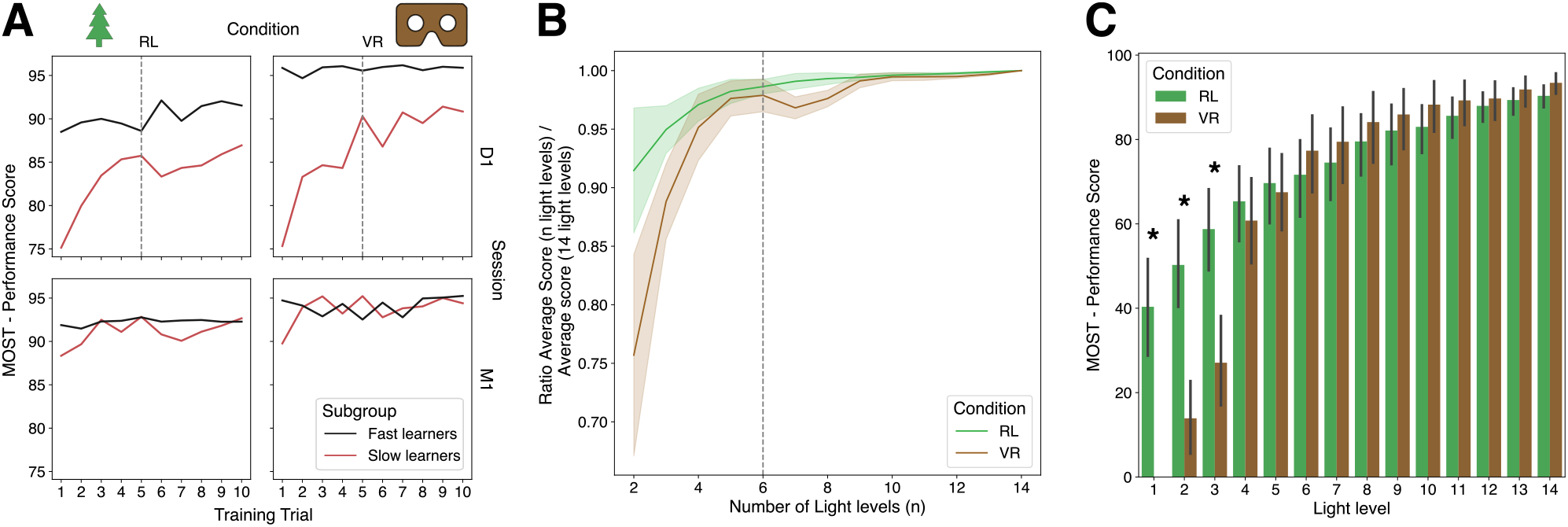
Main experimental results from Phase 2 of the study. **A**: Evolution of the performance score during training for RL (left), VR (right), D1 (day one, top) and M1 (month one, bottom). Patients were divided in two groups: the fast learners (n=25) having few performance improvements during learning, and slow learners (n=5). **B**: Relation between the resampled number of light levels and the ratio between the average performance score with n light levels and with 14 light levels. A unity ratio means that the resampled average performance score resampled is close to the average performance score with all light levels. The dashed vertical line indicates the optimal and minimal number of light levels required to have a good agreement. **C**: Performance score as a function of light level (from low [1] to high [14] light levels). RL and VR conditions are depicted in green and brown, respectively. Within mean and standard deviation are represented, as well as significant differences between VR and RL conditions (*). In **B** and **C**, the performance score was averaged over sessions (D1 & M1).

Patients’ performance in the MOST test decreased sharply as the luminance level decreased (F(13,169)=70.44, p<.001, ηp2=.55), and it was lower in VR than RL (F(1,13)=29.75, p<.001, ηp2=.02), but only for the 3 lowest low light levels (See Figure 2.C, interaction Light*Condition: F(13,169)=27.04, p<.001, ηp2=.19). These results led us to adjust the VR light levels in Phase 3 to be more comparable to the RL luminance levels. Since in clinical trials in ophthalmology, the therapy is administered on a single eye in a first phase, our test has to be performed monocularly and binocularly, which increases the number of experimental conditions and, thus, the test duration. We therefore estimated the minimum number of luminance levels to ensure a reproducible score, by interpolating the performance score for a theoretical number of trials between 2 (only the extreme light levels) and 14 light levels (Figure 2.B). Results showed that 6 light levels were sufficient to achieve an average performance comparable to that averaged across 14 light levels.

### Phase 3 - MOST validation

In a third phase, 60 participants (30 RP and 30 controls) performed 4 MOST sessions: in RL and VR conditions, the first day (D1) and one month after (M1). After 5 training trials with the highest luminance level, they performed 18 test trials, in monocular and binocular conditions, from the lowest to the highest light level (6 levels). All participants were able to complete the test, in both RL and VR conditions. The duration of a session was significantly longer for RP patients in RL (110±27 min.) than in VR (62 ±27 min) (t(29) = -10.7, p < .001). Results include the repeatability of the test between sessions (D1/M1), the reliability between modalities (VR/RL), the effect of light level on performance, the construct and content validity and the subjective assessment of MOST.

### Repeatability and reliability

As shown in Table 2 and Figure 3.A&B, we used the intra-class correlations (ICC), mean difference, and 95% limits of agreement (Bland-Altman) to examine the repeatability of the MOST test between sessions. Results showed little to no learning effect between sessions (D1/M1) in both RL and VR conditions. As this agreement is excellent (ICCs > .98), we will only present the average results between D1 and M1 in the following sections. The agreement between the performance score in RL and VR conditions is also excellent, as demonstrated by significant correlations (Figure 3.C) and ICCs (all > .98; Table 2), thus indicating excellent reliability between test modalities.

**Table 2.**
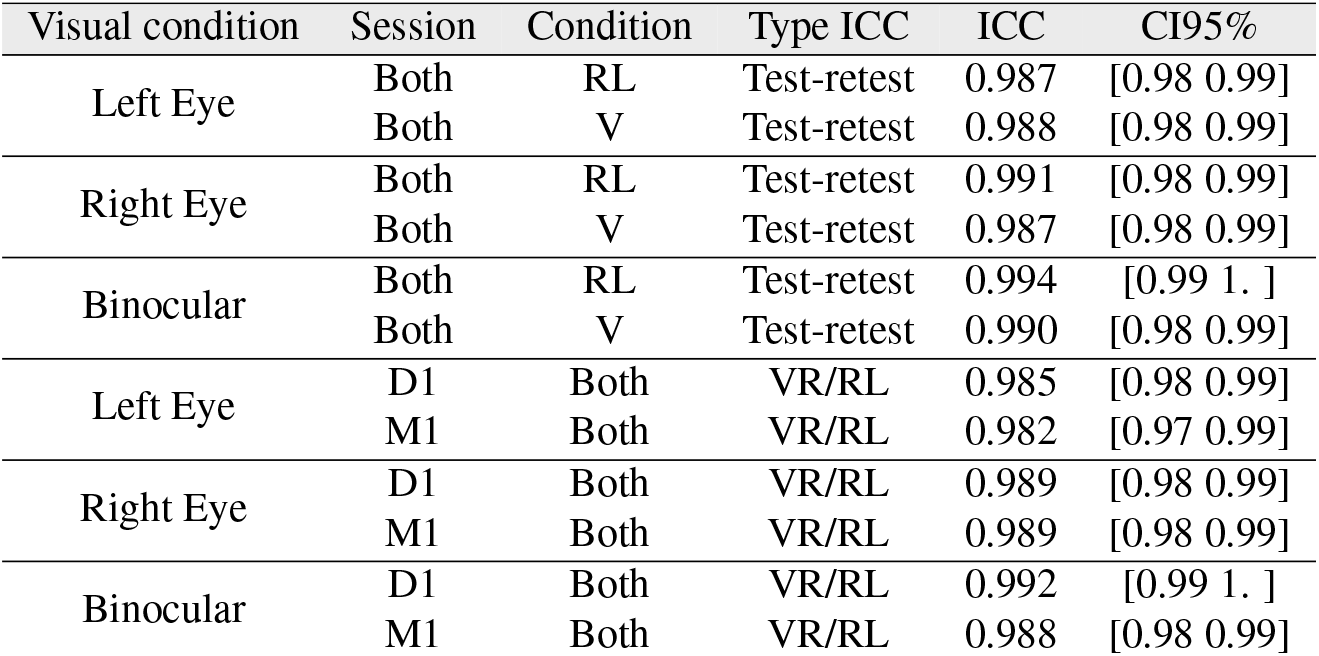
Agreement of the average MOST performance score between session (D1/M1) and between conditions (RL/VR) in Phase 3 of the study. Intra-class correlations (ICC) and confidence intervals (CI95%) are displayed for all visual condition (Left Eye, Right Eye, Binocular).

| Visual condition | Session | Condition | Type ICC | ICC | CI95% |
| --- | --- | --- | --- | --- | --- |
| Left Eye | Both | RL | Test-retest | 0.987 | [0.98 0.99] |
|  | Both | V | Test-retest | 0.988 | [0.98 0.99] |
| Right Eye | Both | RL | Test-retest | 0.991 | [0.98 0.99] |
|  | Both | V | Test-retest | 0.987 | [0.98 0.99] |
| Binocular | Both | RL | Test-retest | 0.994 | [0.99 1. ] |
|  | Both | V | Test-retest | 0.990 | [0.98 0.99] |
| Left Eye | D1 | Both | VR/RL | 0.985 | [0.98 0.99] |
|  | M1 | Both | VR/RL | 0.982 | [0.97 0.99] |
| Right Eye | D1 | Both | VR/RL | 0.989 | [0.98 0.99] |
|  | M1 | Both | VR/RL | 0.989 | [0.98 0.99] |
| Binocular | D1 | Both | VR/RL | 0.992 | [0.99 1. ] |
|  | M1 | Both | VR/RL | 0.988 | [0.98 0.99] |

**Fig. 3.**
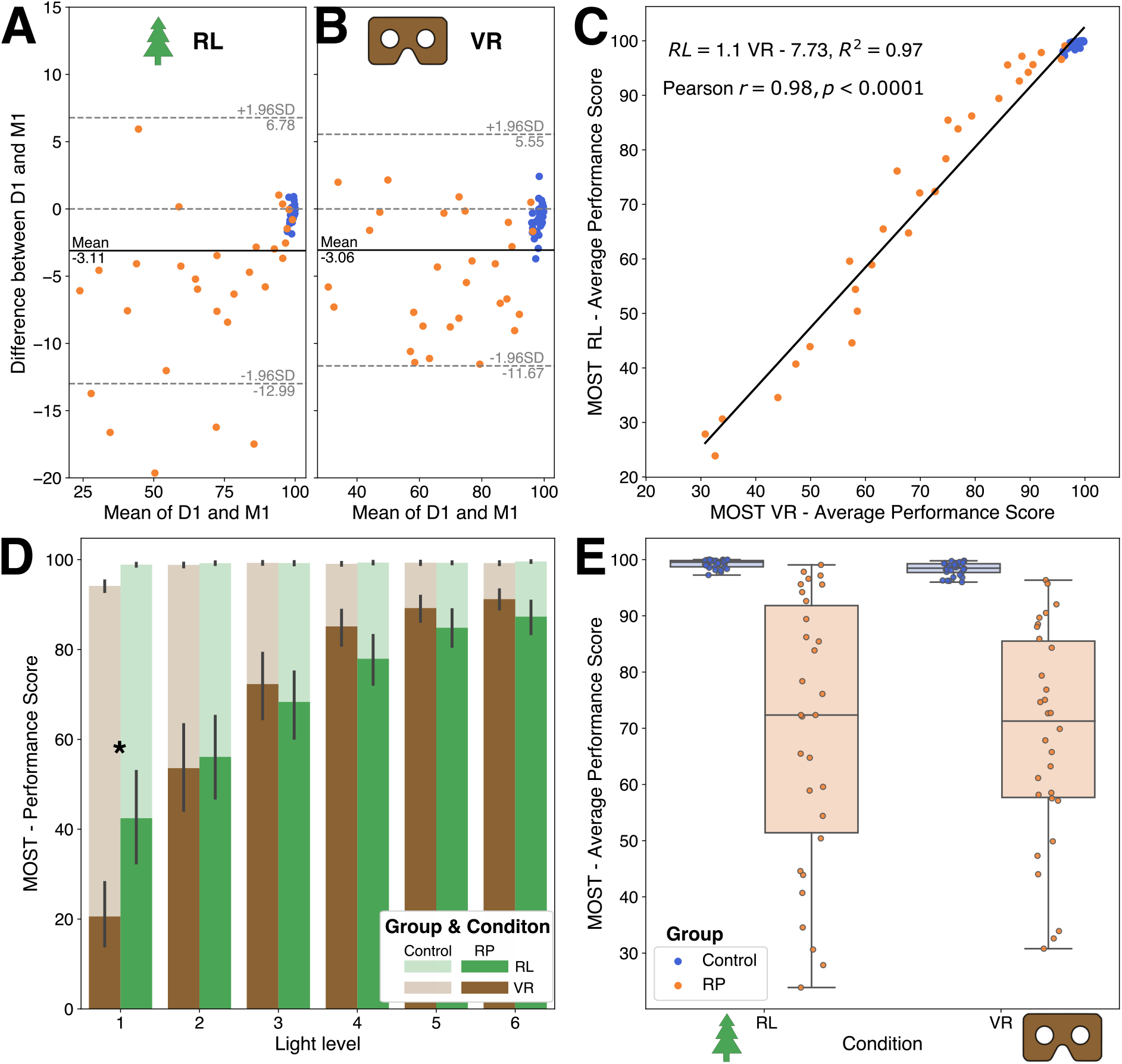
Main experimental results from Phase 3 of the study, in binocular condition. Bland-Altman plots showing the agreement of the average MOST performance score between two sessions (D1/M1) for RL (**A**) and VR (**B**) conditions. The continuous black line represents the average difference between sessions, and the dashed gray lines the limit of agreement. **C**: Correlations between the average physical (RL) and virtual (VR) performance score in binocular condition. **D**: Performance score as a function of light level (from low [1] to high [5] light levels). RL and VR conditions are depicted in green and brown, respectively, as control group is transparent and RP group opaque. Within mean and standard deviation are represented, as well as significant difference between VR and RL conditions (*). **E**: Box plot of the average performance for each group (blue: control; orange: RP) and condition (RL, VR). In **A, B, C** and **E**, each point represents a participant (blue: control; orange: RP). In **C, D** and **E**, the performance score was averaged over sessions (D1 & M1).

### Effect of luminance level for individuals with RP

As in Phase 2, patients’ performances in the MOST test decreased significantly as the light level lowered (F(5,145)=71.13, p<.001, partial ηp2=.71). Overall performances were similar between VR and RL conditions (F(1,29)=0.79, p=.38) although the scores were lower in VR than RL under the lowest luminance condition (See Figure 3.D, interaction Light*Condition: F(5,145)=32.11, p<.001, ηp2=.52).

### Construct validity

We assessed construct validity by characterizing the between-group discriminatory power of the mean performance score. The discrimination ability was close to perfect in all experimental conditions (RL, VR, D1, M1, binocular and monocular conditions), with accuracy, sensitivity and specificity always greater than 95%, 96%, 93%, respectively (Supplementary Table 3). In the worst case scenario, only two RP patients and one control out of 60 participants were misclassified, and those RP patients were in the early stages of the disease (see Figure 3.E). Content validity. Content validity was characterized by testing the ability to discriminate the performance score between different stages of the disease in the RP group(24), and by correlation analyses between the performance score and individuals’ with RP visual variables. Results across conditions revealed a mean 82.29% accuracy, 91.14% sensitivity and 75.77% specificity (Supplementary Table 3). Moreover, as shown in Table 3, the average performance score was also strongly correlated with visual acuity (negative correlation), contrast sensitivity, and visual field measurements: Octopus mean sensitivity, adaptometry thresholds at 5 and 20 minutes, and multiple Goldmann perimetry parameters (see also Supplementary Figure 3 and Supplementary Table 4).

**Table 3.** Relation between binocular MOST performance score and visual characteristics in the RP group (Phase 3). Pearson r and p statistic (corrected for multiple tests) are reported for both RL and VR conditions.

| Visual Variable | RL |  | VR |  |
| --- | --- | --- | --- | --- |
|  | r | p | r | p |
| Visual Acuity (Binocular) | -0.57 | 0.001 | -0.59 | 0.001 |
| Contrast Sensitivity (Binocular) | 0.60 | 0.001 | 0.57 | 0.001 |
| Octopus - Mean Sensitivity (Best Eye) | 0.71 | <0.001 | 0.67 | <0.001 |
| Adaptometry Threshold (5 min., Binocular) | 0.94 | <0.001 | 0.93 | <0.001 |
| Adaptometry Threshold (20 min., Binocular) | 0.91 | <0.001 | 0.90 | <0.001 |
| Goldmann - Central Island Area I4 (Best Eye) | 0.67 | <0.001 | 0.61 | 0.001 |
| Goldmann - Total Area I4 (Best Eye) | 0.79 | <0.001 | 0.75 | <0.001 |
| Goldmann - Central Island Area III4 (Binocular) | 0.60 | 0.001 | 0.58 | 0.001 |
| Goldmann - Total Area III4 (Binocular) | 0.65 | <0.001 | 0.68 | <0.001 |

### Subjective assessment

In Phase 3, the patients completed a questionnaire on the acceptance of the test and VR usability (Supplementary Table 1). Test duration was considered acceptable for a majority of patients (RL: 93%; VR: 97%), who also considered the test pleasant when performed in VR (70%), whereas this rate dropped to 43% in RL. Importantly, a large majority of the patients considered that the difficulties encountered in the test were representative of those encountered in their daily-life (RL: 73%; VR: 80%), and 97% of them suggested using this activity as an assessment of their functional vision abilities. Regarding VR, all participants were able to perform the test without nausea or vertigo, and none of the participants felt that they had put themselves in danger during the test.

## Discussion

In this study, we developed a novel performance-based outcome for evaluating functional vision in inherited retinal diseases, with a focus on RP. The MOST testing paradigm is based on the evaluation of performance in a mobility test, performed in both real and virtual conditions. To validate the MOST, we established a method to control the experimental conditions, in a standardized mobility test as natural as possible, and we quantified participants’ performance with a continuous composite score. The MOST protocol provides control of learning effect within and between sessions (agreement), it is highly correlated between real and virtual reality conditions (fidelity), it is sensitive to disease progression, and it shows a good construct and content validity.

### An outcome in highly controlled experimental conditions

A key aspect in the design of an outcome is the control of experimental biases. In a locomotion test, the most obvious one is the learning of mobility courses. Therefore, the courses must be both sufficiently numerous and comparable in difficulty. Our results showed that MOST’s 28 mobility courses are comparable in difficulty. This was tested in a dedicated experimental phase (Phase 1) unlike other studies(12, 13). We also avoided another bias by controlling for the lighting conditions. This is crucial, because lighting is the physical parameter that is most related to mobility performance in RP patients(15). Luminance levels were carefully controlled in both RL and VR conditions, with very good spatial homogeneity (RL), and a constant log lux-level step between light conditions(27). We also used monochromatic objects to control only the light level in the scene.

### An ecological test, representative of mobility

The main challenge of this study was to design a test that could be performed in both real and virtual conditions. The previously proposed MLMT(12) is performed in a small space (1.6 by 3.1 m), leading to non-natural walking speed (patients: 0.04 m/s; controls: 0.24 m/s, to be compared to a normal walking speed of ∼1.4 m/s(30)). MOST’s locomotion space is larger (5.2 by 3.6 m), which allows for more natural walking speeds (RPs: 0.5 m/s; controls: 0.99 m/s). Moreover, in order to avoid VR-related motion sickness (reported by Lam et al.(17) in glaucoma patients), MOST requires participants to physically move to navigate the virtual environment. Finally, the ecological validity of MOST is endorsed by patient reported outcomes, who consider the difficulties encountered in MOST as representative of those experiences in their daily life (80% in VR condition).

### A continuous scoring system to assess performance

The MLMT assessment introduced an original scoring system(12). It combines two sub-scores – accuracy and duration scores – to determine the ability of the patient under given light condition to pass the test, under ad hoc thresholds. The resulting global score corresponds to the minimum light level passed (from -1: the patient is unable to pass the test at 400 lux, to 6: the patient is able to pass the test at the minimum light level of 1 lux). This approach has the merit to associate the two main variables encountered in mobility tests, and often analyzed separately(14). Indeed, usual variables are mostly either characteristic of the speed at which the task is performed (duration, and walking speed) or accuracy variables (obstacle contacts, deviation from an optimal path). In general, accuracy variables allow normally sighted participants and low-vision patients to be better discriminated with respect to speed variables(31, 32), although this is not always the case(14). Combining accuracy and speed into a single variable makes the results even more predictive(31) of visual disease. Time and accuracy are indeed closely related: the faster a participant moves, the more likely they are to hit obstacles. However, the MLMT score measures only an ability on an ordinal scale, not a continuous performance. The MOST score provides a continuous measure of performance, combining test duration and several accuracy-related variables. We believe that this approach is crucial to increase the sensitivity of the test, to detect changes related to the disease progression – shown by high correlation values between performance and visual variables – or to the effect of a therapy. Control of learning effects within a session. An important aspect of the reproducibility and reliability of an outcome is the control of potential learning effects of all participants during a session, before actually starting the test runs. Indeed, a learning effects would bias the performance outcome as a function of luminance level. The results from the Phase 2 of our study show that 5 trials are sufficient to reach a maximal performance in the task. This is a critical finding, as low-vision patients are more likely to show larger and longer improvements over trials.

### Test-retest agreement

In assessing MOST repeatability, we found an excellent agreement (all ICCs > .98) of the performance score, in all experimental conditions (RL & VR, monocular & binocular) and all groups (RP & controls). This agreement is better than previous studies under real conditions, as in the MLM(12) (correlation between session ∼0.86) and Kumaran et al. (2020(14), repeatability coefficient of 1.10 m/s). Moreover, the small mean difference results in Bland-Altman plots (3%) confirms the measurement stability between sessions. These results indicate that it is not necessary to repeat MOST multiple times before and after an intervention in a clinical trial. This repeatability is even comparable, if not superior, to some tests of visual function, such as the Goldmann perimetry(33) or the dark adaptation test(34). The latter, to be used in clinical studies, should be repeated 5 times before and after treatment to increase repeatability3(35).

### Matching MOST in VR and RL

To the best of our knowledge, the study presented here is the first that compares the mobility performance of visually impaired patients in real and virtual conditions. This achievement was made possible by a crossed design of these two modalities. To date, it is impossible to fully reproduce in VR all physical characteristics of a visual scene (contrast, resolution, light level). Therefore, we first empirically selected the light levels in VR (Phase 2), and we used the patients’ performance, lower in VR than in RL, to re-calibrate the minimum light level in VR. After this calibration, the results of Phase 3 showed that performance was equivalent in RL and VR conditions, as quantified by a significant correlation (r = 0.98). These results demonstrate that MOST-VR is predictive of real-world mobility performance. The use of MOST-VR also has the advantages of being shorter and insensitive to subjective external monitoring (for mobility error measurements), while being safe for participants.

### Sensitivity to categorize stages of the disease

An important aspect of the construct validity of a test is its ability of the test to differentiate clinically distinct groups (e.g., visually impaired vs controls). This requirement was verified by our study, as MOST could successfully discriminate RPs and controls (accuracy larger than 95% in all conditions), even though the RP group included patients in an early phase of the disease. Moreover, content validity was fulfilled by MOST’s ability to discriminate patients at various stages of the disease (mean accuracy of 82.29%), and by the strong correlations between visual function (acuity, contrast sensitivity, dark adaptation, visual field) and functional vision (MOST score). To our knowledge, such strong correlations were never reported in the literature, most notably with adaptometry thresholds (r>0.9, see Table 3 and Supplementary Table 4).

### Limitations

The equipment used for VR had a field of view of 110°, and its pixel density was well below normal human acuity. Because its resolution is lower, this device may not be suitable for tasks in which the influence of peripheral vision is dominant, although resolution in the periphery is lower. Moreover, the mobility test was not fully ecological (a maze with gray objects), although 80% of patients considered it to be representative of difficulties encountered in daily life.

## Conclusion

MOST showed an excellent construct validity, reliability and content validity in both RL and VR conditions. The MOST-VR is suitable and exportable for monitoring the progression of a retinal disease and assessing the efficiency of new treatments. Additional data are awaited to measure the ability of MOST to detect changes such as an improvement or deterioration in the visual condition of a patient (i.e., sensitivity to change), and the adaptive strategies developed by patients(36). Our approach combining real/virtual validation is particularly promising. However, it might not be transferable to all activities of daily life, especially the most ecological ones that cannot be easily reproduced in both VR and RL condition (e.g., street crossing, visual search in a complex and realistic scene). Beyond the applicability to vision impairment assessment, standardized, reproducible and affordable evaluation of therapeutic benefits in clinical trials and post-market, This approach can be tailored to replicate other types of vision performances (e.g., central vision and dexterity) using similar methodologies. The demonstration that VR compares favorably to naturalistic experimental paradigms and that it can meet a higher patient acceptance is promising in terms of development for other visual, neurological, musculoskeletal, and behavioral conditions.

## Data Availability

All data produced in the present study are available upon reasonable request to the authors.

## ACKNOWLEDGEMENTS

The authors would like to express their sincere thanks to the patients who participated in the studies, the Streetlab team that contributed to the acquisition of the data in MOST-RL and MOST-VR (Caroline Kurek, Paul Thomas, Étienne Violain, Chloé Pagot, Christopher Reeves, Charlotte Leflaëc, Julien Adrian, Karine Becker, Emmanuel Gutman); participated to software development (Yihan Zhang, Johan Lebrun, Nathan Flambard, Yichao Liu); and the collection of visual data in collaboration with the XV-XX hospital (Caroline De Montleau, Suzon Ajasse, Darine Marion, Wahiba Khemliche). We would like to warmly thank Catherine Agathos, Angelo Arleo and Daniel Chung for their careful proofreading.

## Appendix 1: Management of light levels in VR and RL conditions

### Physical luminance levels (RL)

The level of lighting in RL condition has been defined according to the number of necessary light conditions (14 in Phase 2, 6 in Phase 3, see Tables 1 and 2). Minimal (1 lux) and maximal (400 lux) light levels were identical between phases, and the other light levels were defined in order to have constant steps in log unit between conditions, as measured with a lux meter (Chroma Meter CL-200A, Konica Minolta, Tokyo, Japan).

**Table 1.** RL light levels in Phase 2

| Light Level<br>Condition | Measurement |  |
| --- | --- | --- |
|  | Lux | Log Lux |
| 1 | 1.00 | 0.00 |
| 2 | 1.59 | 0.20 |
| 3 | 2.51 | 0.40 |
| 4 | 3.99 | 0.60 |
| 5 | 6.32 | 0.80 |
| 6 | 10.02 | 1.00 |
| 7 | 15.88 | 1.20 |
| 8 | 25.18 | 1.40 |
| 9 | 39.93 | 1.60 |
| 10 | 63.30 | 1.80 |
| 11 | 100.37 | 2.00 |
| 12 | 159.13 | 2.20 |
| 13 | 252.29 | 2.40 |
| 14 | 400.00 | 2.60 |

**Table 2.** RL light levels in Phase 3

| Light Level<br>Condition | Measurement |  |
| --- | --- | --- |
|  | Lux | Log Lux |
| 1 | 1.00 | 0.00 |
| 2 | 3.31 | 0.52 |
| 3 | 10.99 | 1.04 |
| 4 | 36.41 | 1.56 |
| 5 | 120.68 | 2.08 |
| 6 | 400.00 | 2.60 |

### Control of luminance levels in VR

The level of lighting in VR was managed in the Unity software used to develop the VR simulation (2018.2.17f1 version in Phase 2, 2019.3.15f1 in Phase 3).

The light in the 3D scene is managed using a combination of several light sources: directional lights and environment lighting (ambient color). Directional lights are used to avoid the uni-color on the objects since if we use only environment light, the different sides of an object will be indistinguishable. We use 4 directional lights without shadows, one on each side of the virtual scene.

Ambient light, also known as diffuse environmental light, is light that is present all around the Scene and doesn’t come from any specific source object. It can be an important contributor to the overall look and brightness of a scene. The environment lighting is introduced to overcome the effect that only the top of an object is visible in dim light condition, since the directional light is projected on the top of the object. We turn off the skybox to avoid potential color added to the scene, and set “Color” as the environment lighting source.

All light sources are controlled with a single parameter in Unity: φ, ranging from 0 (no light) to no upper limit. A value greater than 1 corresponds to a scene that is too bright.

In VR, we chose the light levels empirically:

- the minimal level corresponds to the minimal light condition for which a normally-sighted participant was able to achieve the mobility task after a phase of adaptation to darkness;
- the maximum level corresponds to a light level which visually match the 400 lux condition in the real environment (i.e., the maximum light level in RL), without being too dazzling/glaring.

We then measured the light levels on the Vive Pro Eye screen with the same luxmeter as RL (Chroma Meter CL-200A), in a scene with a mobility course displayed, with the virtual camera positioned on a single position (starting point in the corner - or starting position of each trial) and an orientation of 30° below the horizon. We then measured the relationship between the light level in the Unity scene (φ) from 0 (no light) to 1 and the physical illumination of the screen (in Lux). A polynomial model was sufficient to explain the relationship between φ and the physical illumination (Equation 1).

Physical illumination (Lux) = *aϕ*^3^ + *bϕ*^2^ + *cϕ* + *d*, with a = 9.67 ; b = -2.30 ; c = 0.43 and d = -0.02

In Phase 2, as in RL condition, we made the choice to have constant steps between physical light level (in Log units), between the first visible condition for a control subject (φ = 0.09) to a comfortable visual condition (φ = 0.99, Table 3), by using Equation 1. After an analysis of participants’ performance under low light conditions (Phase 2), we increased the minimum light level for Phase 3 (φ = 0.12, Table 4), as low light levels were too difficult for RP participants in VR condition, as compared to RL condition.

**Table 3.** VR light levels in Phase 2

| Table 3 – VR light levels in Phase 2 |  |  |  |
| --- | --- | --- | --- |
| Light Level<br>Condition | Measurement | | $\varphi$ (Unity) |
|  | Lux | Log Lux |  |
| 1 | 0.01 | -1.89 | 0.10 |
| 2 | 0.02 | -1.67 | 0.13 |
| 3 | 0.03 | -1.46 | 0.16 |
| 4 | 0.06 | -1.25 | 0.20 |
| 5 | 0.09 | -1.03 | 0.24 |
| 6 | 0.15 | -0.82 | 0.29 |
| 7 | 0.25 | -0.61 | 0.34 |
| 8 | 0.40 | -0.39 | 0.40 |
| 9 | 0.66 | -0.18 | 0.47 |
| 10 | 1.08 | 0.03 | 0.54 |
| 11 | 1.76 | 0.25 | 0.63 |
| 12 | 2.88 | 0.46 | 0.73 |
| 13 | 4.71 | 0.67 | 0.85 |
| 14 | 7.70 | 0.89 | 1.00 |

**Table 4.** VR light levels in Phase 3

| Table 4 – VR light levels in Phase 3 |  |  |  |
| --- | --- | --- | --- |
| Light Level<br>Condition | Measurement | | $\varphi$ (Unity) |
|  | Lux | Log Lux |  |
| 1 | 0.02 | -1.71 | 0.12 |
| 2 | 0.06 | -1.19 | 0.21 |
| 3 | 0.21 | -0.67 | 0.32 |
| 4 | 0.70 | -0.15 | 0.47 |
| 5 | 2.33 | 0.37 | 0.69 |
| 6 | 7.70 | 0.89 | 1.00 |

**Supplementary Figure 1.**
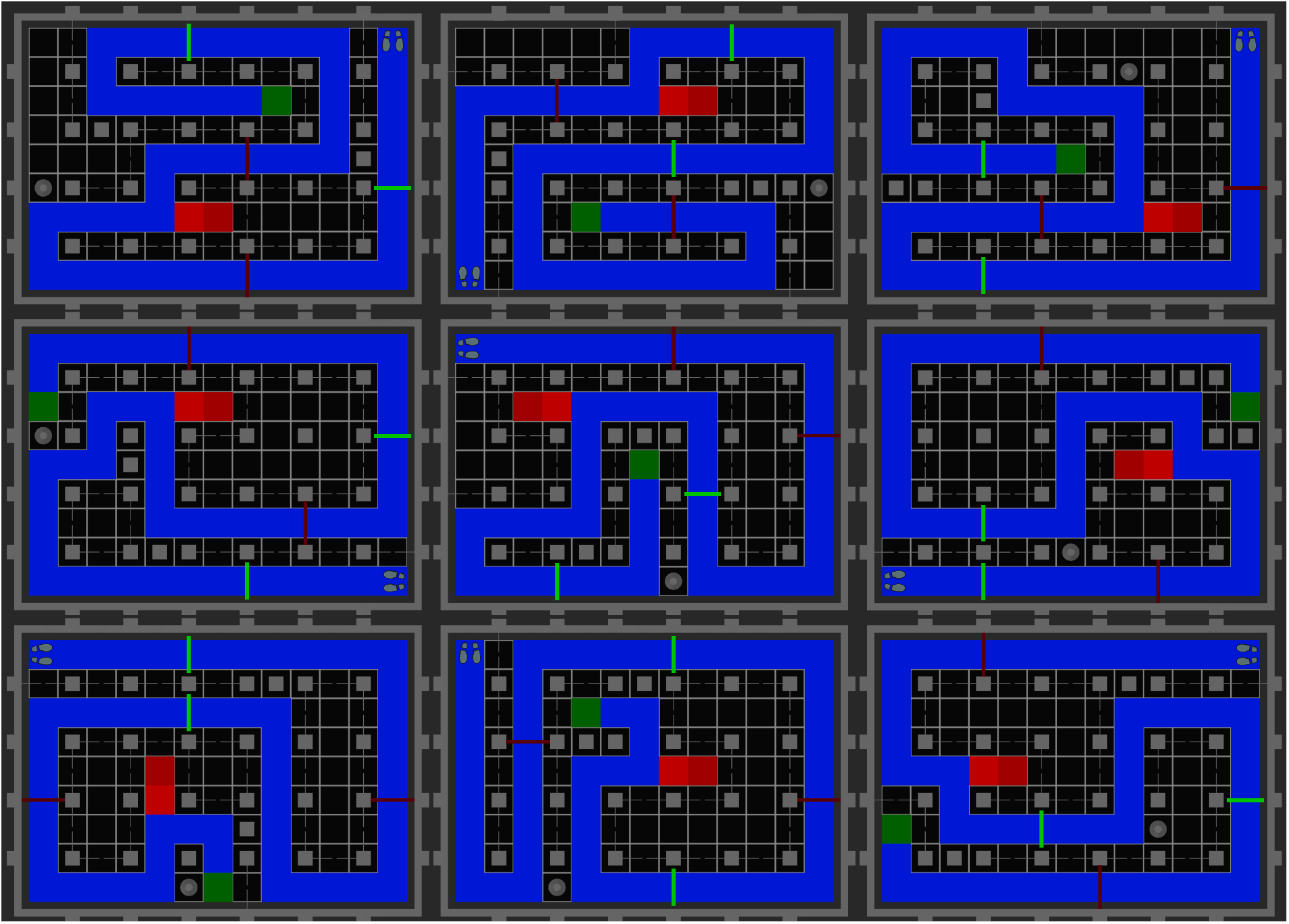
Schematic top view of six mobility courses. The starting point is represented by footprints, the goal as a green square, and the dead end by red tiles. Only one trajectory – displayed in blue on this figure – links the starting point and the destination, as closed doors obstruct the other potential paths. Two types of obstacles are on participant’s path: two flags (red lines) and two steps (green lines). Two other type of objects close the path: a cone and two high columns.

**Supplementary Figure 2.**
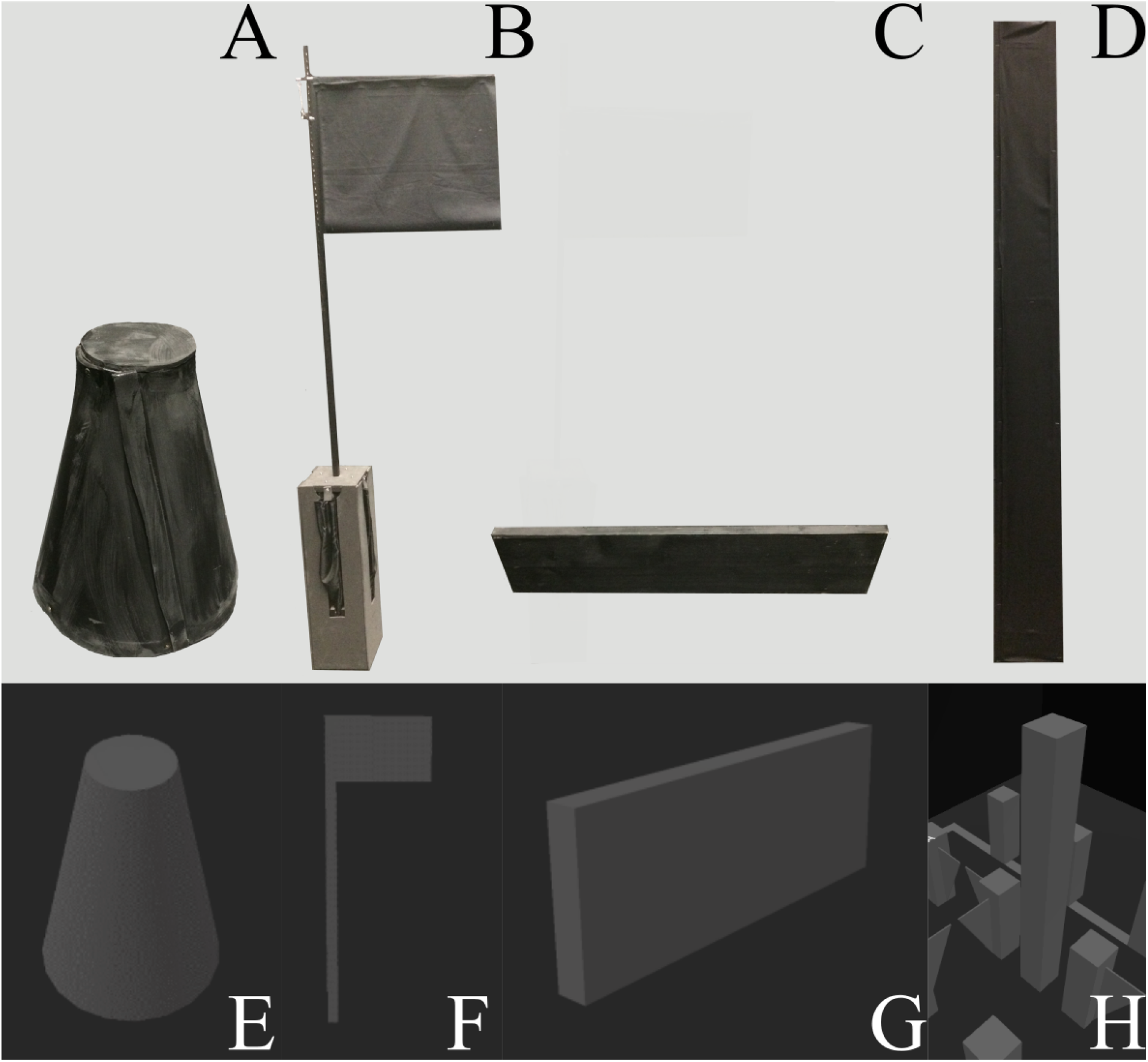
View of the different elements of the course that the participant will encounter (top: RL, bottom: VR). The cone (A/E) and the columns (D/H) close the path, while the steps (C/G) and the flags (B/F) are located on the path, and must be crossed.

**Supplementary Figure 3:**
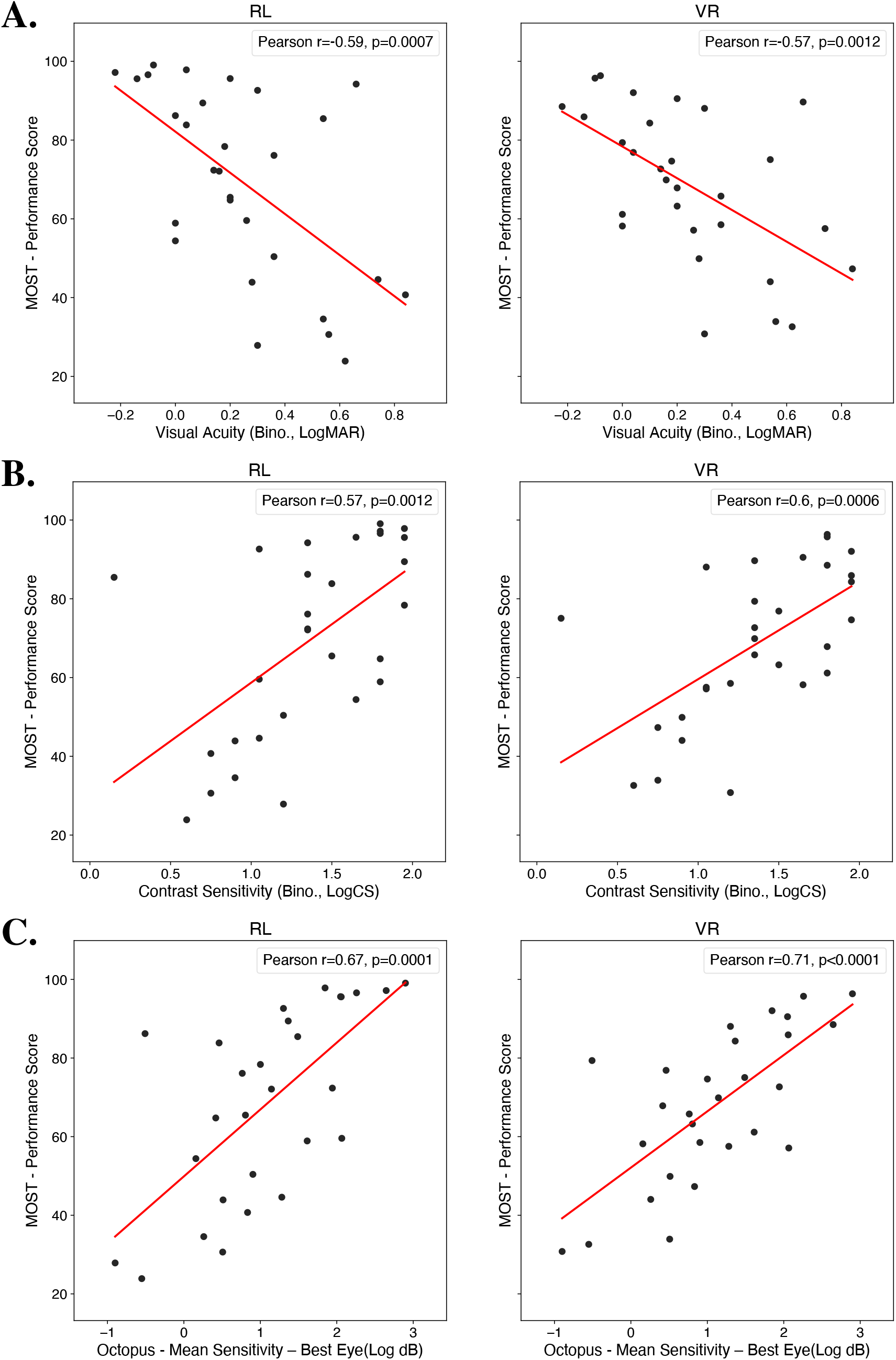
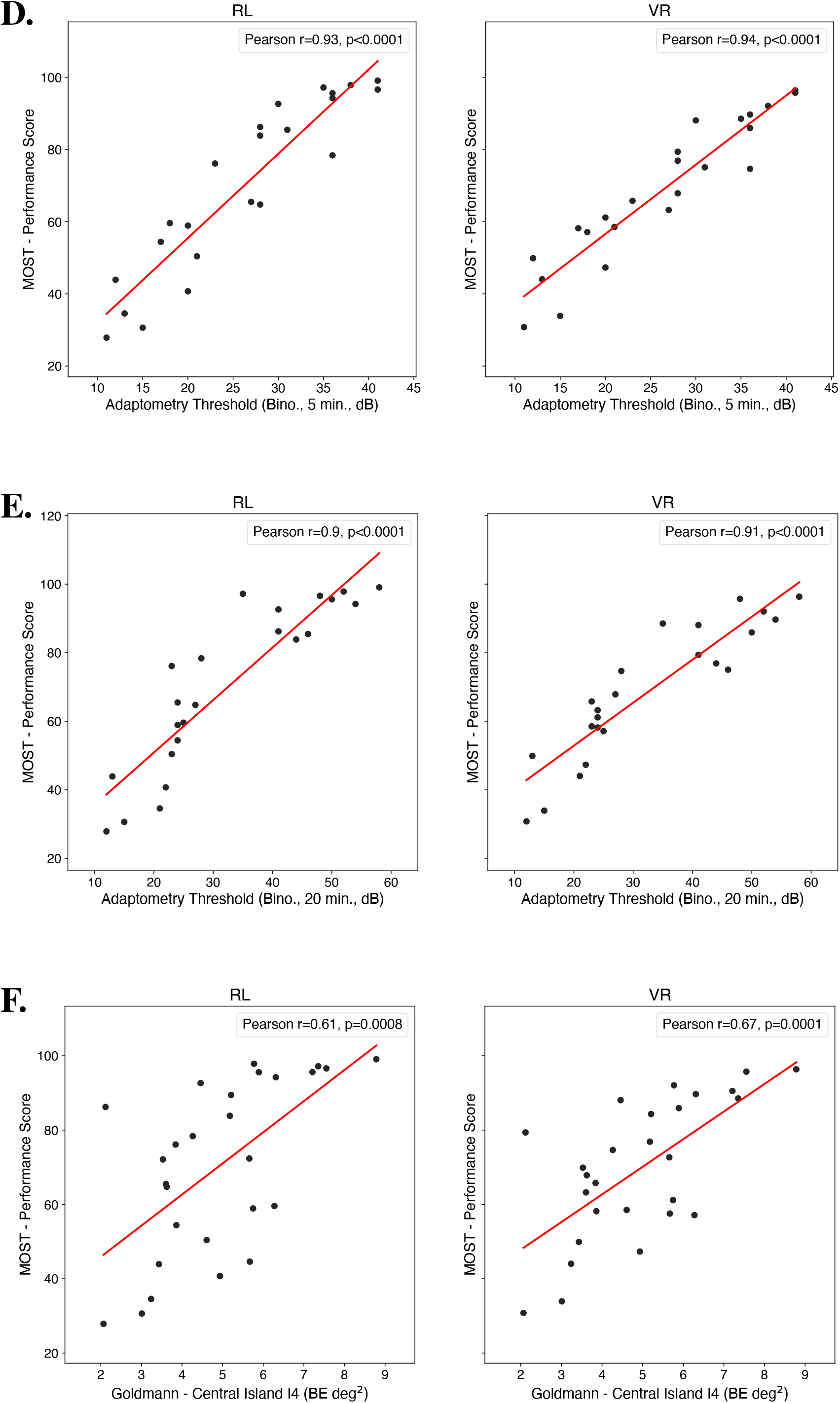
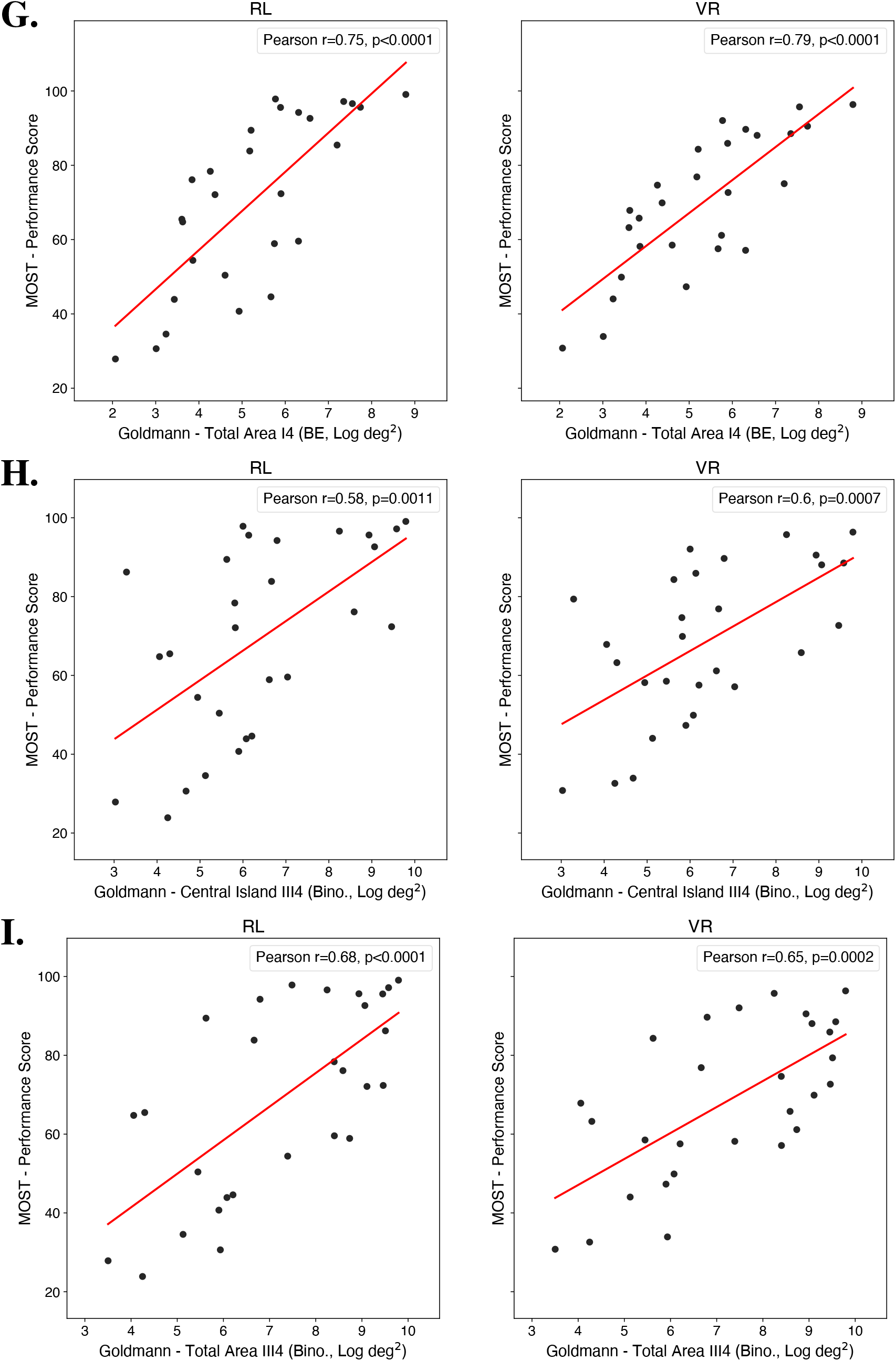
Correlations plots between MOST performance score in binocular condition and visual characteristics (Phase 3). Correlations for both RL (Real Life) and VR (Virtual Reality) are represented. Performance scores are averaged among D1 and M1 sessions. Each point represents an RP participant. Correlations were computed with Visual Acuity (**A**.), Contrast Sensitivity (**B**.), Mean Sensitivity from Octopus (**C**.), Dark Adaptation thresholds (Adaptometry) after 5 (**D**.) and 20 minutes (**E**.), Goldmann central island area with I4 (**F**.) and III4 (**H**.), and Goldmann total area with I4 (**G**.) and III4 (**I**.).

**Supplementary Table 1:**
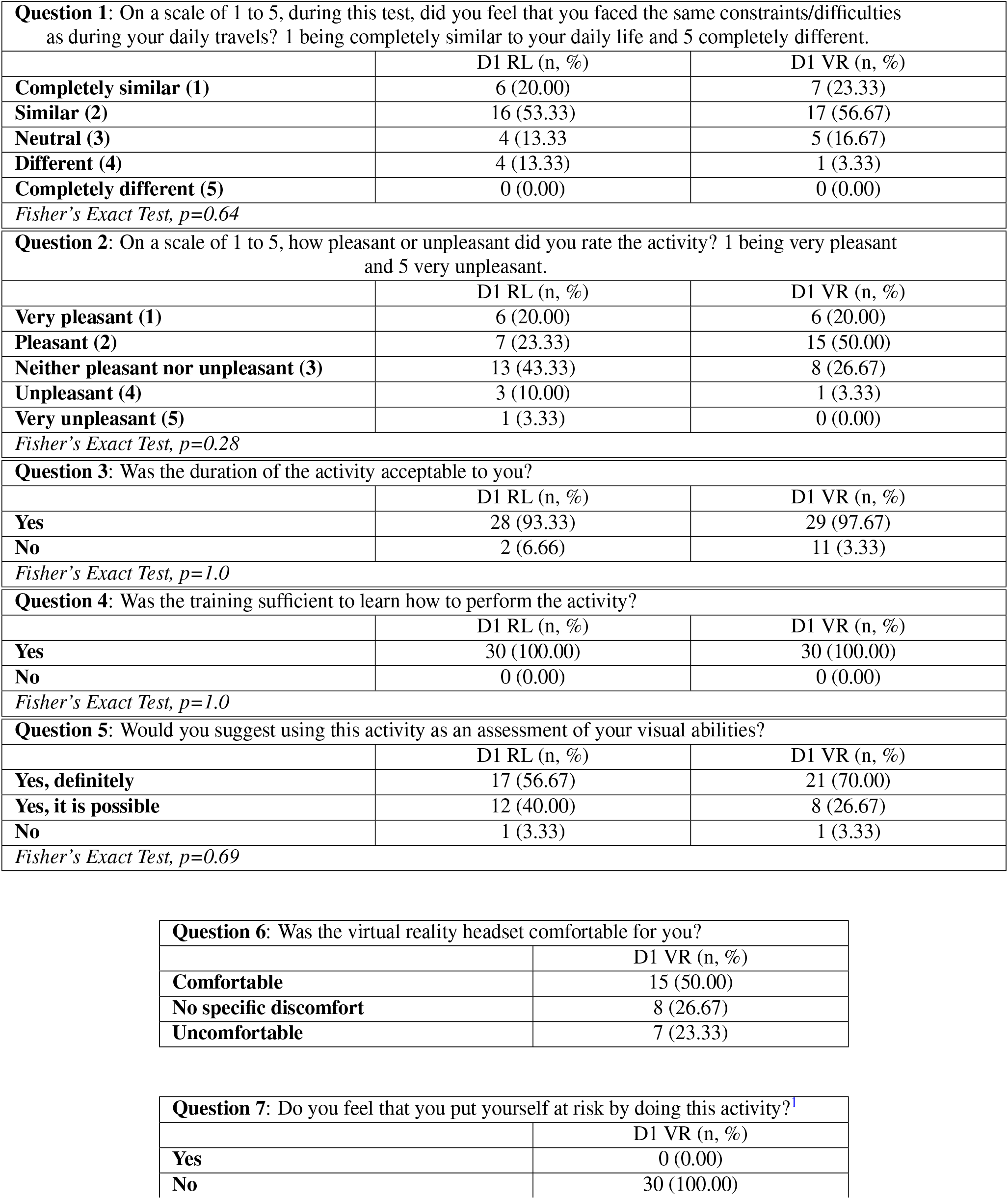
Results of post-test questionnaires completed by RP patients in Phase 3 (N = 30 for all questions). For questions asked in both the real (RL) and virtual (VR) conditions, the statistical value of Fisher’s exact test is shown at the bottom of the table.

|  |  |  |
| --- | --- | --- |
| **Question 1:** On a scale of 1 to 5, during this test, did you feel that you faced the same constraints/difficulties as during your daily travels? 1 being completely similar to your daily life and 5 completely different. |  |  |
|  | D1 RL (n, %) | D1 VR (n, %) |
| **Completely similar (1)** | 6 (20.00) | 7 (23.33) |
| **Similar (2)** | 16 (53.33) | 17 (56.67) |
| **Neutral (3)** | 4 (13.33) | 5 (16.67) |
| **Different (4)** | 4 (13.33) | 1 (3.33) |
| **Completely different (5)** | 0 (0.00) | 0 (0.00) |
| *Fisher's Exact Test, p=0.64* |  |  |
| **Question 2:** On a scale of 1 to 5, how pleasant or unpleasant did you rate the activity? 1 being very pleasant and 5 very unpleasant. |  |  |
|  | D1 RL (n, %) | D1 VR (n, %) |
| **Very pleasant (1)** | 6 (20.00) | 6 (20.00) |
| **Pleasant (2)** | 7 (23.33) | 15 (50.00) |
| **Neither pleasant nor unpleasant (3)** | 13 (43.33) | 8 (26.67) |
| **Unpleasant (4)** | 3 (10.00) | 1 (3.33) |
| **Very unpleasant (5)** | 1 (3.33) | 0 (0.00) |
| *Fisher's Exact Test, p=0.28* |  |  |
| **Question 3:** Was the duration of the activity acceptable to you? |  |  |
|  | D1 RL (n, %) | D1 VR (n, %) |
| **Yes** | 28 (93.33) | 29 (97.67) |
| **No** | 2 (6.66) | 11 (3.33) |
| *Fisher's Exact Test, p=1.0* |  |  |
| **Question 4:** Was the training sufficient to learn how to perform the activity? |  |  |
|  | D1 RL (n, %) | D1 VR (n, %) |
| **Yes** | 30 (100.00) | 30 (100.00) |
| **No** | 0 (0.00) | 0 (0.00) |
| *Fisher's Exact Test, p=1.0* |  |  |
| **Question 5:** Would you suggest using this activity as an assessment of your visual abilities? |  |  |
|  | D1 RL (n, %) | D1 VR (n, %) |
| **Yes, definitely** | 17 (56.67) | 21 (70.00) |
| **Yes, it is possible** | 12 (40.00) | 8 (26.67) |
| **No** | 1 (3.33) | 1 (3.33) |
| *Fisher's Exact Test, p=0.69* |  |  |

|  |  |
| --- | --- |
| **Question 6:** Was the virtual reality headset comfortable for you? |  |
|  | D1 VR (n, %) |
| **Comfortable** | 15 (50.00) |
| **No specific discomfort** | 8 (26.67) |
| **Uncomfortable** | 7 (23.33) |
| **Question 7:** Do you feel that you put yourself at risk by doing this activity?<sup>1</sup> |  |
|  | D1 VR (n, %) |
| **Yes** | 0 (0.00) |
| **No** | 30 (100.00) |

**Supplementary Table 2.** Effect of mobility course configurations on mean participant performance in Phase 1: duration of the trial (in seconds) and number of collisions (± standard deviation), number of interventions, obstacle errors (flag and step) and entrance to the dead end. The last line of the table indicate the p-value of the repeated-measure ANOVA, showing no difference of performance between mobility courses.

| Mobility course # | Duration (s) | Collisions | Interventions | Flag | Step | Dead end |
| --- | --- | --- | --- | --- | --- | --- |
| 1 | 17.44 (2.66) | 0.20 (0.41) | 0 | 0 | 0 | 0 |
| 2 | 17.70 (2.77) | 0.07 (0.26) | 0 | 0 | 0 | 0 |
| 3 | 18.12 (3.12) | 0.00 (0.00) | 0 | 0 | 0 | 0 |
| 4 | 17.51 (2.97) | 0.07 (0.26) | 0 | 0 | 0 | 0 |
| 5 | 18.16 (3.56) | 0.07 (0.26) | 0 | 0 | 0 | 0 |
| 6 | 18.18 (2.90) | 0.33 (0.49) | 0 | 0 | 0 | 0 |
| 7 | 17.93 (2.61) | 0.07 (0.26) | 0 | 0 | 0 | 0 |
| 8 | 17.83 (2.64) | 0.00 (0.00) | 0 | 0 | 0 | 0 |
| 9 | 17.57 (2.34) | 0.27 (0.46) | 0 | 0 | 0 | 0 |
| 10 | 17.67 (2.48) | 0.13 (0.35) | 0 | 0 | 0 | 0 |
| 11 | 17.68 (2.64) | 0.00 (0.00) | 0 | 0 | 0 | 0 |
| 12 | 17.51 (3.13) | 0.00 (0.00) | 0 | 0 | 0 | 0 |
| 13 | 17.91 (3.26) | 0.20 (0.56) | 0 | 0 | 0 | 0 |
| 14 | 18.18 (3.72) | 0.07 (0.26) | 0 | 0 | 0 | 0 |
| 15 | 17.94 (2.54) | 0.13 (0.35) | 0 | 0 | 0 | 0 |
| 16 | 17.78 (3.09) | 0.13 (0.35) | 0 | 0 | 0 | 0 |
| 17 | 17.84 (2.47) | 0.07 (0.26) | 0 | 0 | 0 | 0 |
| 18 | 17.56 (2.58) | 0.13 (0.35) | 0 | 0 | 0 | 0 |
| 19 | 17.56 (2.55) | 0.00 (0.00) | 0 | 0 | 0 | 0.07 |
| 20 | 17.26 (2.09) | 0.27 (0.70) | 0 | 0 | 0 | 0 |
| 21 | 17.56 (3.13) | 0.07 (0.26) | 0 | 0 | 0 | 0 |
| 22 | 17.67 (2.47) | 0.27 (0.59) | 0 | 0 | 0 | 0 |
| 23 | 17.76 (2.49) | 0.00 (0.00) | 0 | 0 | 0 | 0 |
| 24 | 18.03 (2.75) | 0.13 (0.35) | 0 | 0 | 0 | 0 |
| 25 | 17.55 (2.62) | 0.13 (0.52) | 0 | 0 | 0 | 0 |
| 26 | 17.75 (2.91) | 0.00 (0.00) | 0 | 0 | 0 | 0 |
| 27 | 17.69 (2.61) | 0.13 (0.35) | 0 | 0 | 0 | 0 |
| 28 | 17.60 (2.46) | 0.13 (0.35) | 0 | 0 | 0 | 0 |
| Anova p-value | .62 | .21 | - | - | - | .46 |

**Supplementary Table 3.** Power of discrimination of the MOST performance score in Phase 3 of the study. The table presents accuracy, sensitivity and specificity (in %) of the classification based on the performance score in MOST. Each experimental condition is analyzed separately: left, right and binocular conditions, and VR and RL conditions. Table present discrimination between groups (RP, control) as well as discrimination between subgroups of RP (early, advanced). The cut-off is determined using Youden’s index.

| Comparison | Visual Condition | Session | Condition | Accuracy | Sensitivity | Specificity |
| --- | --- | --- | --- | --- | --- | --- |
| Between Groups (RP/Control) | Left Eye | D1 | RL | 98.33 | 100.00 | 96.77 |
|  |  |  | VR | 98.33 | 100.00 | 96.77 |
|  |  | M1 | RL | 98.33 | 100.00 | 96.77 |
|  |  |  | VR | 98.33 | 100.00 | 96.77 |
|  | Right Eye | D1 | RL | 96.67 | 96.67 | 96.67 |
|  |  |  | VR | 96.67 | 100.00 | 93.75 |
|  |  | M1 | RL | 98.33 | 100.00 | 96.77 |
|  |  |  | VR | 96.67 | 100.00 | 93.75 |
|  | Binocular | D1 | RL | 95.00 | 96.55 | 93.55 |
|  |  |  | VR | 96.67 | 100.00 | 93.75 |
|  |  | M1 | RL | 96.67 | 96.67 | 96.67 |
|  |  |  | VR | 98.33 | 100.00 | 96.77 |
| Between RP subgroups (early/advanced) | Left Eye | D1 | RL | 83.33 | 100.00 | 73.33 |
|  |  |  | VR | 79.17 | 100.00 | 68.75 |
|  |  | M1 | RL | 79.17 | 83.33 | 75.00 |
|  |  |  | VR | 79.17 | 83.33 | 75.00 |
|  | Right Eye | D1 | RL | 87.50 | 91.67 | 83.33 |
|  |  |  | VR | 83.33 | 90.91 | 76.92 |
|  |  | M1 | RL | 83.33 | 90.91 | 76.92 |
|  |  |  | VR | 87.50 | 91.67 | 83.33 |
|  | Binocular | D1 | RL | 83.33 | 90.91 | 76.92 |
|  |  |  | VR | 83.33 | 90.91 | 76.92 |
|  |  | M1 | RL | 79.17 | 90.00 | 71.43 |
|  |  |  | VR | 79.17 | 90.00 | 71.43 |

**Supplementary Table 4.** Relation between monocular MOST performance score and monocular visual characteristics in the RP group (Phase 3 of the study). Pearson r and p statistic (corrected for multiple tests) are reported for both RL and VR conditions.

| Visual Variable | RL |  | VR |  |
| --- | --- | --- | --- | --- |
|  | r | p | r | p |
| Visual Acuity (Left Eye) | -0.52 | 0.005 | -0.57 | 0.002 |
| Contrast Sensitivity (Left Eye) | 0.49 | 0.009 | 0.48 | 0.009 |
| Octopus - Mean Sensitivity (Left Eye) | 0.83 | <0.001 | 0.79 | <0.001 |
| Goldmann - Central Island Area I4 (Left Eye) | 0.65 | 0.001 | 0.60 | 0.002 |
| Goldmann - Total Area I4 (Left Eye) | 0.74 | <0.001 | 0.70 | <0.001 |
| Goldmann - Central Island Area III4 (Left Eye) | 0.54 | 0.004 | 0.47 | 0.011 |
| Goldmann - Total Area III4 (Left Eye) | 0.66 | <0.001 | 0.64 | 0.001 |
| Visual Variable | RL |  | VR |  |
|  | r | p | r | p |
| Visual Acuity (Right Eye) | -0.56 | 0.002 | -0.56 | 0.002 |
| Contrast Sensitivity (Right Eye) | 0.61 | 0.001 | 0.59 | 0.001 |
| Octopus - Mean Sensitivity (Right Eye) | 0.80 | <0.001 | 0.77 | <0.001 |
| Goldmann - Central Island Area I4 (Right Eye) | 0.80 | <0.001 | 0.79 | <0.001 |
| Goldmann - Total Area I4 (Right Eye) | 0.82 | <0.001 | 0.81 | <0.001 |
| Goldmann - Central Island Area III4 (Right Eye) | 0.64 | <0.001 | 0.60 | 0.001 |
| Goldmann - Total Area III4 (Right Eye) | 0.61 | 0.001 | 0.60 | 0.001 |

